# Clinical Application of Coronary Computed Tomography Angiography in Patients with Suspected Coronary Artery Disease

**DOI:** 10.1101/2022.06.23.22276823

**Authors:** Jared N. Blad, Hayden L. Smith, Jonathan R. Hurdelbrink, Steven R. Craig, Brent Wolford, Elizabeth Wendl, Nelson A. Telles-Garcia

## Abstract

**Background:** Coronary computed tomography angiography (CCTA) has emerged as a helpful tool in the evaluation of patients with chest pain and suspected coronary artery disease (CAD). The study objective was to evaluate the clinical application of CCTA for assessment of patients presenting with chest pain at a large single urban tertiary health care center located in the US.

**Methods:** A retrospective review of observational data from center was conducted. The study sample included outpatients aged 18 years or older who underwent CCTA during January 2018 through February 2020.

**Results:** There were 158 patients in the study, with sixty-two percent having stress testing within 6-months prior to CCTA. Fifty-five percent of these patients had abnormal stress test findings but demonstrated no CAD on CCTA. Among the patients with normal stress tests, 24.4% demonstrated CAD on CCTA. Twenty-five patients underwent invasive coronary angiography (ICA) within the 6-months following CCTA. For these patients, the positive predictive value of CCTA was 91.3% (95% CI: 72.0%, 99.0%) and negative predictive value was 100.0% (95% CI: 15.8%, 100.0%) when compared to ICA results. In addition, CCTA demonstrated myocardial bridging in 10.0% and an anomalous coronary vessel in 8.8% of patients.

**Conclusions:** This study revealed that CCTA is being used as a diagnostic test in patients presenting with chest pain at the study center. Among patients with prior stress testing, these results were at times potentially inconsistent with the findings from the CCTA. In addition, the CCTA was able to identify variants in anatomical structures.

## Introduction

An initial diagnostic approach for patients with suspected coronary artery disease (CAD) often consists of stress testing. The type of stress test utilized is dependent upon patient characteristics, pre-test probability, and expertise available in the geographic area. Exercise electrocardiogram (ECG) is the most widely available and least expensive functional test, but it has limited diagnostic power in-regards to CAD.^1,2^ Other stress test modalities use echocardiography or gamma-nuclear imaging to investigate for myocardial ischemia. Stress testing is a physiological test which assesses ischemia which is a surrogate for anatomic coronary stenosis.

Coronary computed tomography angiography (CCTA) is an emerging diagnostic tool for evaluation of patients presenting with symptoms suggestive of cardiac ischemia. The PROMISE, SCOT-HEART, and EVINCI studies suggested that CCTA improved diagnostic confidence and supported the use of CCTA for evaluation of patients with chest pain.^3-5^ As a result, the National Institute for Health and Care Excellence recommends CCTA as a first-line approach for all patients presenting with chest pain.^6^ The recent American College of Cardiology (ACC) and American Heart Association (AHA) guidelines for the diagnosis and management of patients with stable chest pain now recommend CCTA as a first-line diagnostic test alternative.^7^ Of note, CCTA has been shown to possibly overestimate the angiographic severity of CAD, although its negative predictive value allows it to effectively exclude significant CAD.^8^

The objective of this study was to assess the clinical application and accuracy of CCTA in a retrospective sample of outpatients at a single tertiary care medical center located in the central United States.

## Methods

### Study Type/Inclusion/Exclusion

A retrospective review of medical record data from a tertiary care, independent academic medical center, was conducted for the 26-month period of January 2018 through February 2020. The sample included patients 18 years or older who underwent CCTA at the study institution for the outpatient workup of suspected CAD. Patients that underwent computed tomography (CT) of the heart for pre-operative assessment prior to aortic valve replacement or ablation were excluded. CCTA studies with non-interpretable results due to poor image quality and studies in which the coronary arteries were not assessed were excluded.

### Data Collection

The electronic medical records for study patients were reviewed to determine CCTA results and potential subsequent invasive coronary angiography (ICA) results. Records were also reviewed to identify any documented subsequent major adverse cardiovascular events (MACE), vascular stenting, and coronary artery bypass grafting (CABG). Patient CAD risk factors were identified, including documented hypertension, hyperlipidemia, obesity, diabetes mellitus, family history of premature CAD, and history of tobacco use. Dates and results of cardiac stress testing and imaging within 6-months prior to and after the CCTA date were also collected.

### Definitions of Measures

Obstructive CAD was defined as greater than 50% documented stenosis of the left main coronary artery or greater than 70% documented stenosis in any other coronary artery as reported in the CCTA or ICA report; these thresholds are comparable to standard delineations of flow-limiting stenosis.^9^ Vessels that were graded as ‘severe’ were also categorized as obstructive.

Non-obstructive CAD was defined as between 20% and 50% documented stenosis of the left main coronary artery or between 20% and 70% in any other coronary artery. Vessels graded as ‘minimal,’ (i.e., stenosis between 1% and 24%) were classified as non-obstructive CAD unless the CCTA or ICA report explicitly noted a percent narrowing of less than 20%. For the left main coronary artery, vessels graded as ‘mild’ (i.e., stenosis between 1% and 24%) or ‘moderate’ (i.e., stenosis between 25% and 49%) were graded as non-obstructive CAD. All other coronary arteries graded as ‘mild’ (i.e., stenosis between 25% and 49%) and ‘moderate’ (i.e., stenosis between 50% and 69%) were also categorized as non-obstructive CAD. No apparent CAD was defined as vessels with luminal irregularities of less than 20% stenosis in all coronary arteries.

Patients who underwent ICA within 6-months after the CCTA were used to calculate the positive and negative predictive values of CCTA. A result of non-obstructive or obstructive CAD on CCTA/ICA was considered a positive result while no apparent CAD was considered a negative CCTA/ICA result.

### CT Procedure

The CCTA scans were obtained on a 64-slice Siemens (Erlangen, Germany) Somatom Definition AS using helical multi-detector computed tomography MDCT acquisition with retrospective ECG-gating, rotation time of 330 milliseconds, collimation of 64 × 0.6 mm, and dose reduction technique.

### Statistical Analysis

Categorical variables were reported as counts with percentages and continuous variables were reported as means with standard deviations (SD). The number of risk factors between the patient obstruction groups were compared using Poisson regression and reported as relative rates. Positive and negative predictive values were calculated to examine testing modalities. Estimates are reported with 95% confidence intervals (CI).

## Results

### Study Participants

An electronic medical record review initially revealed 208 patients underwent CCTA during the 26-month study period. After the application of the study exclusion criteria (Figure 1), there were 158 patients included in the final sample. The mean patient age was 49.9 (SD: 16.1) years, with 88.0% of the sample being white and 56.3% being female. Of these patients, 17.1% had a history of diabetes mellitus, 52.5% were obese, 50.6% hypertensive, 50.0% had hyperlipidemia, 39.2% had a current or prior history of tobacco use with 25.3% having a family history of premature CAD. Myocardial bridging of a coronary vessel was documented in 10.0% (n=17) of patients and 8.8% (n=15) of patients were reported as having an anomalous origin or malignant course of a large epicardial coronary vessel per CCTA. Of these patients, an anomalous aortic origin of a coronary vessel was documented in 60.0% (n=9), Figure 2.

**Figure 1:**
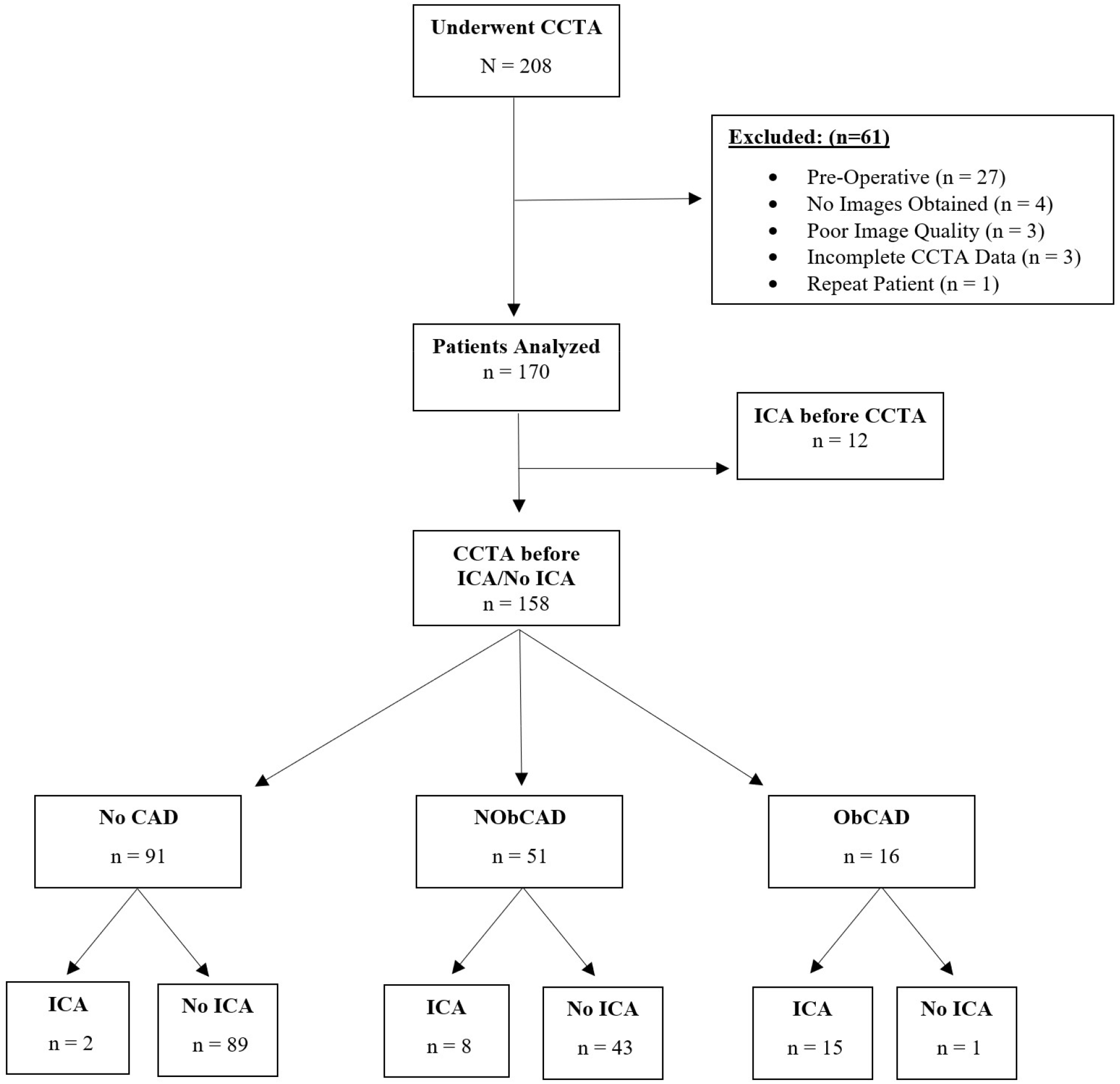
Flowchart of the application of inclusion/exclusion criteria for a retrospective observational study conducted over a 26-month period from January 2018 to February 2020 at an independent academic medical center in the United States. CCTA: Coronary computed tomography angiography; ICA: Invasive coronary angiography; CAD: Coronary artery disease; NObCAD: Non-obstructive coronary artery disease; ObCAD: Obstructive coronary artery disease.

**Figure 2:**
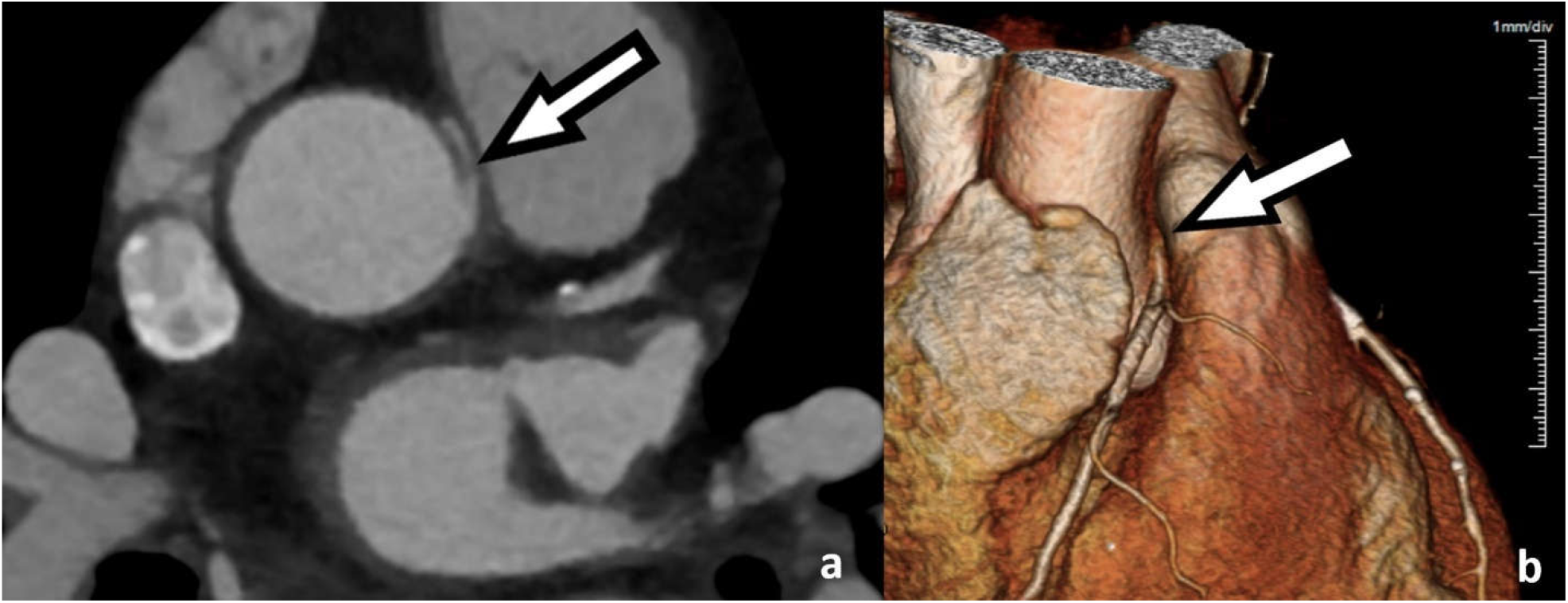
Two and three dimensional coronary computed tomography angiography images demonstrating a right coronary artery (RCA) that arises from the left coronary cusp with slit like origin and acute angle at the takeoff (a) as well as inter-arterial, so-called malignant, course. The RCA can be visualized emerging between the aorta and the pulmonary trunk (b).

### CCTA Results

Fifty-eight percent (n=91) of the 158 patients were found to have no apparent CAD, 32.2% (n=51) had non-obstructive CAD and 10.1% (n=16) had obstructive CAD on CCTA testing. A total of 25 of the 158 patients underwent ICA within 6-months following CCTA. Of patients with a CCTA result of obstructive CAD, 93.8% (n=15) underwent subsequent ICA, while 2.2% (n=2) of patients with no apparent CAD and 15.7% (n=8) of patients with non-obstructive CAD underwent subsequent ICA (Figure 3). In the 158 patients, the mean number of documented cardiac risk factors was 2.4 out of 6. Patients with CCTA results of no apparent CAD had a mean of 1.8 risk factors while patients with non-obstructive CAD and obstructive CAD had means of 3.0 and 3.3, respectively (Table 1). These differences were represented by a 1.7 (95% CI: 1.3, 2.1) times greater rate for non-obstructive and 1.8 (95% CI: 1.2, 2.5) times greater rate for obstructive CAD for risk factors than those with no apparent CAD on CCTA.

**Figure 3:**
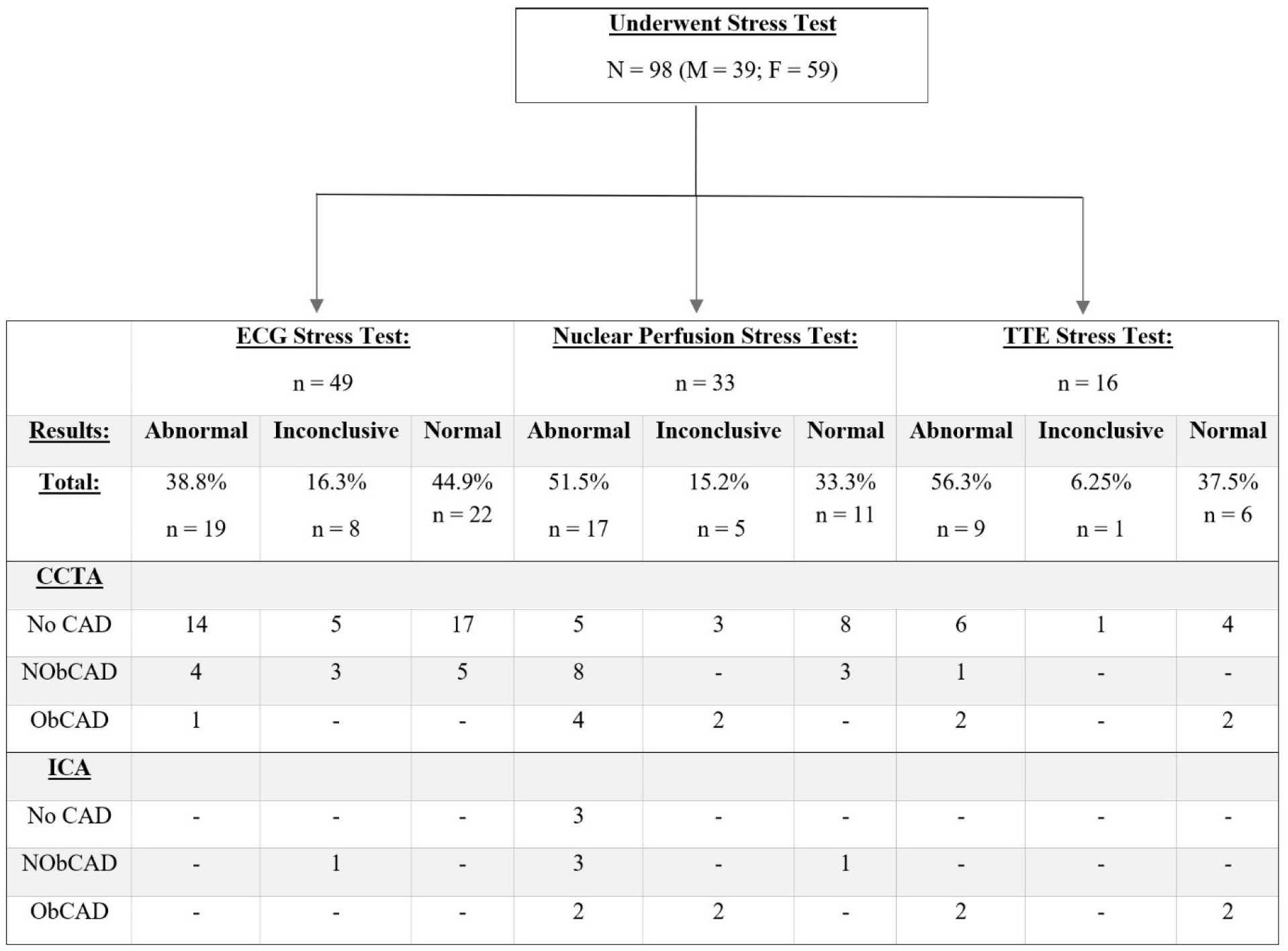
Flow-table from a retrospective observational study conducted over a 26-month period from January 2018 to February 2020 at an independent academic medical center in the United States comparing results of ECG Stress tests, nuclear perfusion stress tests and transthoracic echocardiogram Stress tests to coronary computed tomography angiography results. CCTA: Coronary computed tomography angiography; CAD: Coronary artery disease; NObCAD: Non-obstructive coronary artery disease; ObCAD: Obstructive coronary artery disease; ICA: Invasive coronary angiography; TTE: Transthoracic echocardiogram.

**Table 1:** Results from a retrospective observational review of patients who underwent CCTA from January 2018 to February 2020 at an independent academic medical center in the Midwest.

|  |  | <u>Full Sample</u> |  | <u>No CAD</u> |  | <u>NObCAD</u> |  | <u>ObCAD</u> |
| --- | --- | --- | --- | --- | --- | --- | --- | --- |
| <u>Variable</u> | <u>N</u> | <u>Mean (Std)</u> | <u>n</u> | <u>Mean (Std)</u> | <u>n</u> | <u>Mean (Std)</u> | <u>n</u> | <u>Mean (Std)</u> |
| Female | 158 | 56.3% (0.5) | 91 | 63.7% (0.48) | 51 | 47.1% (0.50) | 16 | 43.8% (0.51) |
| Age | 158 | 49.9 (16.11) | 91 | 42.6 (14.89) | 51 | 59.9 (11.01) | 16 | 59.7 (14.63) |
| Diabetes | 158 | 17.1% (0.38) | 91 | 12.1% (0.33) | 51 | 23.5% (0.43) | 16 | 25.0% (0.45) |
| Hypertension | 158 | 50.6% (0.50) | 91 | 39.6% (0.49) | 51 | 62.8% (0.49) | 16 | 75.0% (0.45) |
| Obesity | 158 | 52.5% (0.50) | 91 | 46.2% (0.50) | 51 | 64.7% (0.48) | 16 | 50.0% (0.52) |
| Hyperlipidemia | 158 | 50.0% (0.50) | 91 | 33.0% (0.47) | 51 | 68.6% (0.47) | 16 | 87.5% (0.34) |
| Family History of CAD | 158 | 25.3% (0.44) | 91 | 18.7% (0.39) | 51 | 33.3% (0.48) | 16 | 37.5% (0.50) |
| Tobacco | 158 | 39.2% (0.49) | 91 | 31.9% (0.47) | 51 | 47.1% (0.50) | 16 | 56.3% (0.51) |
| Underwent Stress Test | 158 | 62.0% (0.49) | 91 | 69.2% (0.46) | 51 | 47.1% (0.50) | 16 | 68.8% (0.48) |
| CCTA prior to Stress Test<sup>a</sup> | 98 | 5.1% (0.22) | 63 | 3.2% (0.18) | 24 | 12.5% (0.34) | 11 | 0.0% (0.00) |
| Stress Test: Abnormal<sup>a</sup> | 98 | 45.9% (0.50) | 63 | 39.7% (0.49) | 24 | 54.2% (0.51) | 11 | 63.6% (0.50) |
| Stress Test: Inconclusive<sup>a</sup> | 98 | 12.2% (0.33) | 63 | 11.1% (0.32) | 24 | 12.5% (0.34) | 11 | 18.2% (0.40) |
| Stress Test: Normal<sup>a</sup> | 98 | 39.8% (0.49) | 63 | 46.0% (0.50) | 24 | 33.3% (0.48) | 11 | 18.2% (0.40) |
| Underwent ICA | 158 | 15.8% (0.37) | 91 | 2.2% (0.15) | 51 | 15.7% (0.37) | 16 | 93.8% (0.25) |
| ICA – No CAD<sup>b</sup> | 25 | 16.0% (0.37) | 2 | 100% (0.00) | 8 | 12.5% (0.35) | 15 | 6.7% (0.26) |
| ICA – NObCAD<sup>b</sup> | 25 | 40.0% (0.50) | 2 | 0.0% (0.00) | 8 | 62.5% (0.52) | 15 | 33.3% (0.49) |
| ICA – ObCAD<sup>b</sup> | 25 | 44.0% (0.51) | 2 | 0.0% (0.00) | 8 | 25.0% (0.46) | 15 | 60.0% (0.51) |
| Underwent Vascular Stenting | 158 | 3.2% (0.18) | 91 | 0.0% (0.00) | 51 | 2.0% (0.14) | 16 | 25.0% (0.45) |
| Underwent CABG | 158 | 1.9% (0.14) | 91 | 0.0% (0.00) | 51 | 2.0% (0.14) | 16 | 12.5% (0.34) |

CCTA: Coronary computed tomography angiography; ICA: Invasive coronary angiography; CAD: Coronary artery disease; NObCAD: Non-obstructive coronary artery disease; ObCAD: Obstructive coronary artery disease; CABG: Coronary artery bypass grafting
a: N= 98 for these variables as only 98 (62.0% of the full 158) underwent any stress test
b: N= 25 for these variables as only 25 (15.8% of the full 158) underwent an ICA

### Testing Sequence

A review of the sequence of tests showed that 62.0% (n=98) of patients underwent stress testing within 6-months of CCTA. Regarding the testing modality, 50.0% (n=49) of the tests were stress ECGs, 33.7% (n=33) were stress nuclear perfusion studies, and 16.3% (n=16) were stress transthoracic echocardiograms (Figure 3). It was found that 45.9% (n=45) of all stress test results were abnormal, 14.3% (n=14) were inconclusive, and 39.8% (n=39) were normal. Of patients that underwent stress testing, only 5.1% (n=5) underwent CCTA prior to the stress test. Of patients with abnormal stress tests, 60.0% (n=18) of females and 46.7% (n=7) of males demonstrated no CAD on CCTA representing a nonsignificant 13.3% (95% CI: -15.7%, 44.1%) rate difference between the sexes. No CAD was found on CCTA in 74% (n=14) of patients that had an abnormal stress ECG (Figure 3). Of patients with normal stress tests, 24.4% (n=10) demonstrated CAD on CCTA (Figure 4). Non-obstructive CAD was found on CCTA in 19.5% (n=8) and obstructive CAD was found on CCTA in 4.9% (n=2) of these patients.

**Figure 4:**
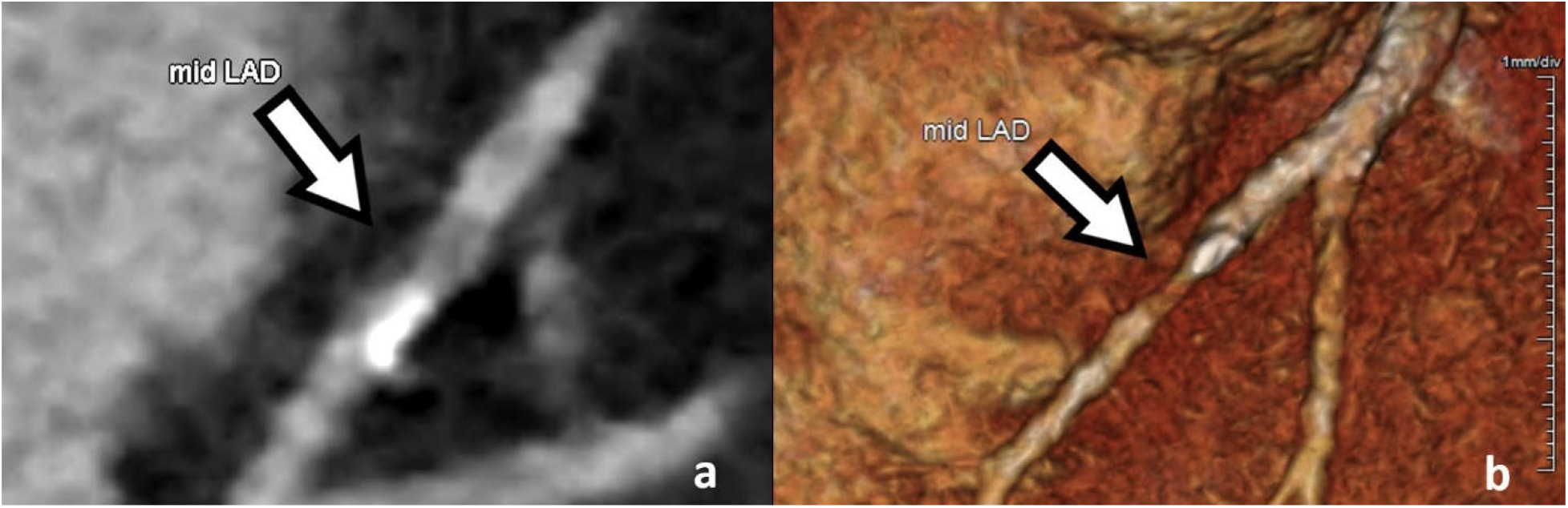
Two dimensional (a) and three dimensional (b) coronary computed tomography angiography imaging of the mid left anterior descending (LAD) artery demonstrating moderate atherosclerotic calcification with moderate to high-grade stenosis in a symptomatic patient who previously had a non-diagnostic treadmill electrocardiogram and a normal myocardial perfusion scan. This patient subsequently underwent coronary angiography which demonstrated a 90% stenosis in the mid LAD requiring stent placement.

### Subsequent Procedures

Following CCTA, 3.1% (n=5) of patients underwent percutaneous coronary intervention (PCI) vascular stenting. Obstructive disease was documented in the CCTA report of 4 patients that underwent subsequent vascular stenting. Non-obstructive disease was documented in 1 patient (Figure 5). Three of these patients had a stress test within 6 months of the CCTA and two did not. Stress test results were inconclusive for 2 patients and were found to be normal for 1 patient. Two percent (n=3) of patients underwent CABG within 6-months following CCTA. Two of these patients had obstructive CAD on CCTA and one had non-obstructive CAD (Figure 5).

**Figure 5:**
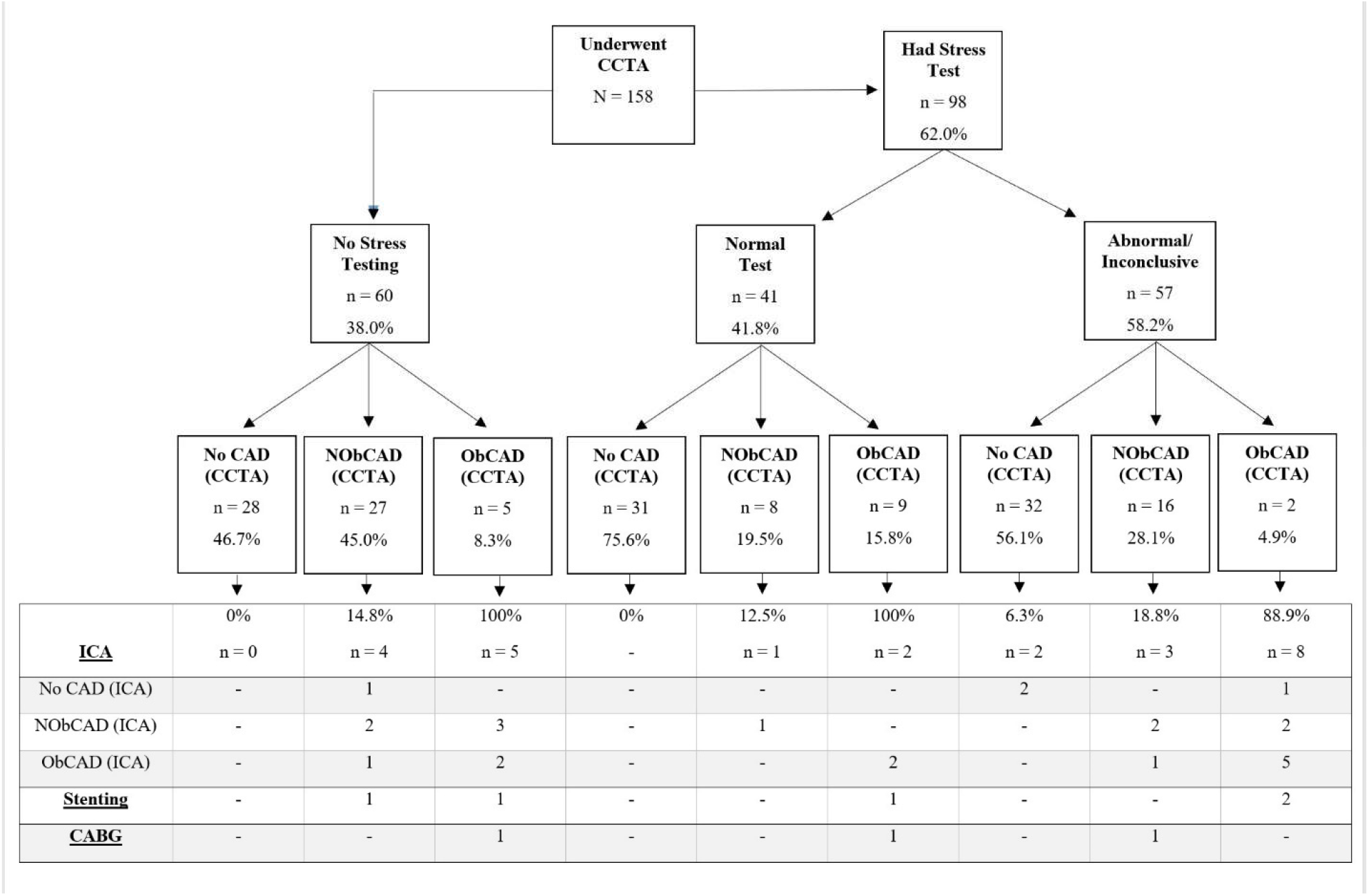
Flow-table compares coronary computed tomography angiography results and subsequent procedures by stress test results from a retrospective observational study conducted over a 26-month period from January 2018 to February 2020 at an independent academic medical center in the United States. CCTA: Coronary computed tomography angiography; CAD: Coronary artery disease; NObCAD: Non-obstructive coronary artery disease; ObCAD: Obstructive coronary artery disease.

The patient with non-obstructive CAD on CCTA underwent CABG to repair an anomalous vessel in which the right coronary artery arose from the left aortic sinus. This patient previously had an abnormal stress test. One other patient that underwent CABG had a stress test within 6 months prior to CCTA. This patient was documented to have had a normal stress test. There were no documented MACE events within the 6-months following the CCTA scan in the full study sample.

### Predictive Value

Twenty-five patients underwent ICA within 6-months following CCTA. The mean time between the two tests was 12.8 (SD: 4.8) days. The positive predictive value of the CCTA was 91.3% (95% CI: 72.0%, 99.0%) and the negative predictive value was 100.0% (95% CI: 15.8%, 100.0%) compared to the ICA.

## Discussion

This retrospective observational study assessed the clinical application of CCTA in the outpatient evaluation of patients with chest pain at a single center for a 26-month period. It was found that over half of patients underwent stress testing within 6-months prior to CCTA. Results suggested that stress testing was not the most reliable predictor of CAD severity in reviewed patients, as approximately half of the patients with abnormal stress tests demonstrated no CAD on CCTA. Further, 4.9% of patients with normal stress tests were found to have severe obstructive CAD on CCTA. One of these patients with a normal stress test required vascular stenting and the other required CABG within the follow up period. Although the sample size was small, the positive predictive value and negative predictive values were consistent with published data on the accuracy of CCTA and may support the potential reliability of CCTA in the assessment of CAD.^3,4^ This would suggest that adopting the National Institute for Health and Care Excellence guidelines by utilizing CCTA earlier in the workup of chest pain may improve diagnostic certainty and treatment efficiency.

Occasionally, CCTA also revealed unexpected findings such as anomalous coronary arteries and myocardial bridging. The prevalence of myocardial bridging has been shown to differ based on the modality of detection.^9^ Rates of myocardial bridging from autopsy series range from 5-86% with a mean of 25%.^10^ A recent study including 4500 patients who had undergone CCTA found the prevalence of myocardial bridging to be 10%.^11^ The present study detected bridging in 17 patients yielding a rate of 10%. The prevalence of anomalous coronary arteries in the general population has been reported to be 1.5% as detected by magnet resonance imaging and 2.6% as detected by CCTA.^12,13^ The present study found that 8.8% of patients that underwent CCTA were documented to have an anomalous vessel and most of these patients were documented to have an anomalous aortic origin of a coronary vessel. The contribution of these findings to chest pain is not entirely clear; however, utilizing CCTA early in the workup of chest pain can aid in identifying non-CAD causes of chest pain and may alter the course of treatment for these patients.

Overall, CCTA appears to be effective in the detection of CAD in the early evaluation of patients presenting with stable chest pain. However, there are certain limitations and challenges that need to be taken into consideration when choosing this imaging technique. According to several meta-analyses, high density calcification can produce blooming artifacts in CCTAs leading to the overestimation of the degree of coronary stenosis and resulting in low positive predictive values.^14-18^ Also, there is a lack of uniform criteria for the assessment of coronary arteries with 50-60% lumen stenosis, which can lead to different interpretation of data.^19,20^ Finally, radiation doses associated with CCTA have gone up in recent years, increasing concerns for related risks.^21^

### Limitations

Given the retrospective study design not all patients who had a CCTA received a subsequent ICA. An ICA was generally performed based on the cardiologist’s interpretation of the clinical presentation, symptoms, and results of other cardiac testing. The present study only included patients undergoing CCTA testing, so patients initially receiving a stress test with no subsequent CCTA would not be captured in the sample. The short follow-up period limited the assessment of outcomes in patients that received CCTA. If patients presented to another institution for a MACE or revascularization, these events may not be included in the patient records. Lastly, the study only reviewed the practices of a single institution, potentially limiting the generalizability of findings.

## Conclusion

This study revealed that CCTA is being used as a first-or second-line diagnostic test in patients presenting with chest pain at the study center. Among patients with prior stress testing, these results were at times potentially inconsistent with findings from the CCTA. In addition, the CCTA was able to identify variants in anatomical structures. The incorporation of CCTA earlier in the evaluation of patients presenting with chest pain should be considered to avoid unnecessary invasive procedures and additional functional tests. Future research should include all patients presenting with chest pain and follow them for a longer time period to fully assess all possible outcomes. Lastly, expanding the sample to include a larger volume of patients from other institutions would more accurately assess the role of cardiac stress testing and CCTA in the evaluation of patients with chest pain.

## Data Availability

All data produced in the present study are not available upon request to the authors

